# A pilot randomised prospective comparison of two approaches for tibial nailing using clinical and novel imaging outcome measures – study protocol

**DOI:** 10.1101/2021.11.18.21266340

**Authors:** Benjamin M Davies, Erden Ali, Daud Chou, Peter Hull, Jaikirty Rawal, James McKay, Andrew McCaskie, Andrew Carrothers

**Author notes:** **Declarations of Interest**. BMD reports funding from an NIHR Clinical Lectureship, not directly related to this study. EA has nothing else to declare. DC has nothing else to declare. PH has nothing else to declare. JW has nothing else to declare. JMcK reports grants and/or consultancy payments from GlaxoSmithKline, Moximed, GE Healthcare, and employment by AstraZeneca, none directly related to this study. AMcC reports work for Taylor and Francis (publishers), membership of an advisory board for Arthritis Research UK, and a patent for cartilage repair unrelated to this study. AC reports grants and/or consultancy payments from NIHR, Stryker, Matortho, Zimmer Biomet, as well as income from expert testimony, none directly related to this study.

## Abstract

Fractures of the tibia are frequently treated with an intra-medullary nail. This can be inserted through either a supra-patella or infra-patella surgical incision. Concerns over potential damage to the cartilage of the knee with supra-patella insertion has impacted upon its adoption despite benefits in terms of easier nail placement and potentially reduced blood loss and pain.

This randomised pilot study will use patient reported outcome measures (PROMS) and novel MRI sequences designed to assess damage to the structure of cartilage to compare these two methods of tibial intra-medullary nailing.

Twenty individuals with tibial shaft fractures will be randomised to either method of nail insertion (ten individuals in each arm). MRI scans and PROMS will be used to assess cartilage damage and general function up to 6 months post operatively.

The results of the study will be used to inform a potential multi-centre study.

## 1 BACKGROUND

Intra-medullary nailing is a common form of treatment for a variety of tibial shaft fractures. Traditionally this has been done via an infra-patella approach. More recently the use of a supra-patella approach has become more popular. However, there are concerns that the supra-patella approach may cause unacceptable damage to the cartilage of the patello-femoral joint (PFJ).

**Figure 1:**
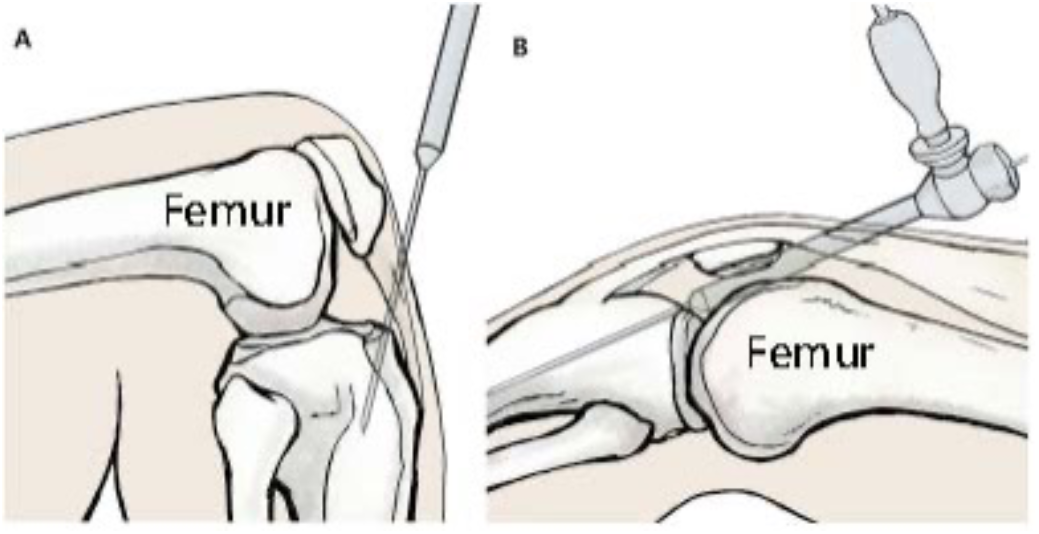
Diagrammatic representations of the infra-patella (A) and supra-patella (B) approaches. Image adapted from Wang et al^14^.

A recent meta-analysis of RCTs indicated that the supra-patella approach may be superior in terms of post-operative pain and blood loss^1^. Unfortunately, none of the four included RCTs included cartilage assessment. Three previous studies have assessed cartilage using standard MRI sequences that demonstrate structural features^2–4^. These were able to show some cases of damage to the cartilage but due to the nature of the MRI sequences used they could not fully evaluate this. Additionally, MRI scanning was not compared to the infra-patella approach.

Three previous cadaveric studies have compared the two different methods to assess cartilage damage. Two studies relied on visual assessment of cartilage damage, and although some cartilage damage was demonstrated this was not statistically or clinically significant^5,6^. The third study measured pressure within the PFJ during intra-medullary nailing^7^. This demonstrated increased pressure in the supra-patella group, but it was not possible to relate this to clinical outcomes given the cadaveric nature of the study.

This study will use a novel MRI technique developed at the University of Cambridge to compare the amount of damage caused to the knee joint by both infra- and supra-patella nailing techniques. Specific cartilage assessment sequences will enable a fuller assessment of cartilage damage than can be achieved with normal structural sequences. Participants will also be asked to complete Patient Reported Outcome Measures (PROMs) regarding their injury to assess their clinical outcome.

Infra-patella and supra-patella nailing techniques are now both routine methods of carrying out intra-medullary nailing of the tibia. Further evidence is required, however, in order to fully assess whether one method is superior to the other.

**Table 1:**
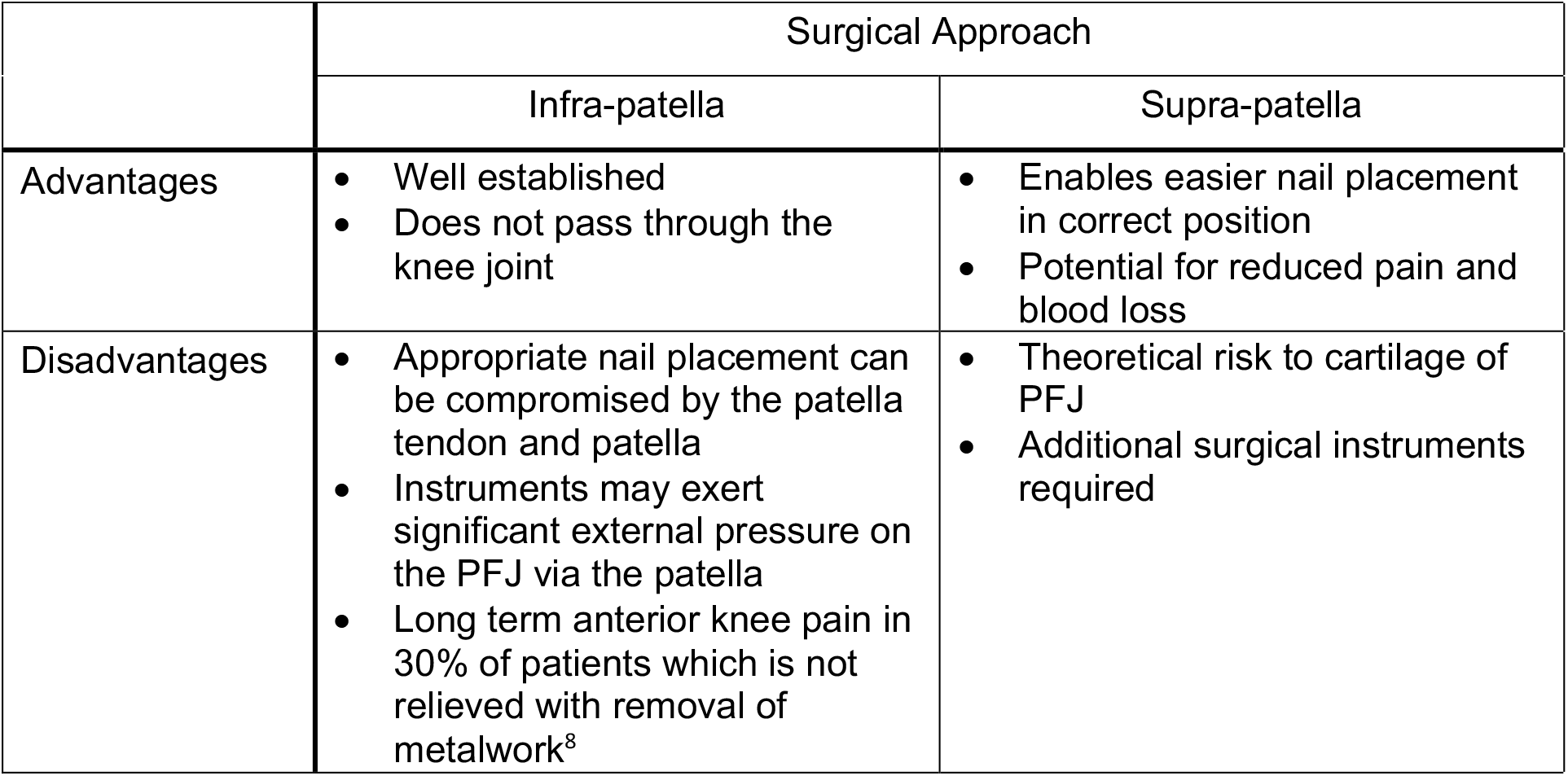
Advantages and Disadvantages of both Surgical Approaches

This study will provide preliminary information to answer this question and generate the data required to support a future nationwide multi-centre study. It will also provide information to support the translation of this technique into the study of other musculoskeletal conditions such as osteoarthritis and intra-articular fractures.

## 2 RATIONALE

This study will provide preliminary information to answer the question of which method of nail insertion causes less damage to the cartilage of the knee. It will generate the data required to support a future nationwide multi-centre study. It will also provide information to support the translation of the novel MRI techniques being employed into the study of other musculoskeletal conditions such as osteoarthritis and intra-articular fractures.

## 3 RESEARCH QUESTION/AIM(S)

### 3.1 Objectives

#### Primary Objective

To determine if there is any difference between infra-patella and supra-patella nailing techniques on immediate and medium-term cartilage structure.

#### Secondary Objective

To determine if there is any difference in clinical outcomes between infra-patella and supra-patella nailing techniques.

### 3.2 Outcome

The aim of the study is to determine if the use of a supra-patella entry point technique (compared with an infra-patella entry point) has any effect upon the degree of cartilage damage to the knee during tibial nailing, both in the short and medium term.

## 4 STUDY DESIGN and METHODS of DATA COLLECTION AND DATA ANALYIS

The study will be a randomised, controlled trial comparing two different methods of inserting a tibial intra-medullary nail.

The two methods of inserting the nail (supra-patella and infra-patella) are both recognised as standard surgical techniques and described in the operative technique manual supplied by the nail manufacturer.

Individuals who are enrolled in the study (see section 6 for participant criteria) will be randomised to one of the two methods. Both of these methods are in routine clinical use at CUHNHSFT. Participants will receive normal care in terms of surgical implants, post-operative rehabilitation, and post-operative follow up. In addition, they will undergo three MRI scans (one pre-operatively and two post operatively) and be asked to complete patient reported outcome measures when they attend for their routine follow up appointments.

Data will be extracted by the clinical care team looking after the patient and then stored in a linked-anonymous format. The data will be held on CUH computers in an encrypted format. Radiology data will be held on CUH radiology systems.

Plain radiographs will be reviewed and interpreted by members of the orthopaedic care team. MRI images will be reviewed and interpreted by Dr J MacKay, a member of the clinical radiology team.

Changes in MRI scores will be compared using ANOVA.

Differences in PROMS scores will be compared using Unpaired T-tests and Mann-Whitney tests (depending on whether the data is normally distributed or not).

Differences in clinical features between the two groups will be compared using Unpaired T-tests and Mann-Whitney tests for continuous data and Fisher’s Exact test for categorical data.

**Figure 2:**
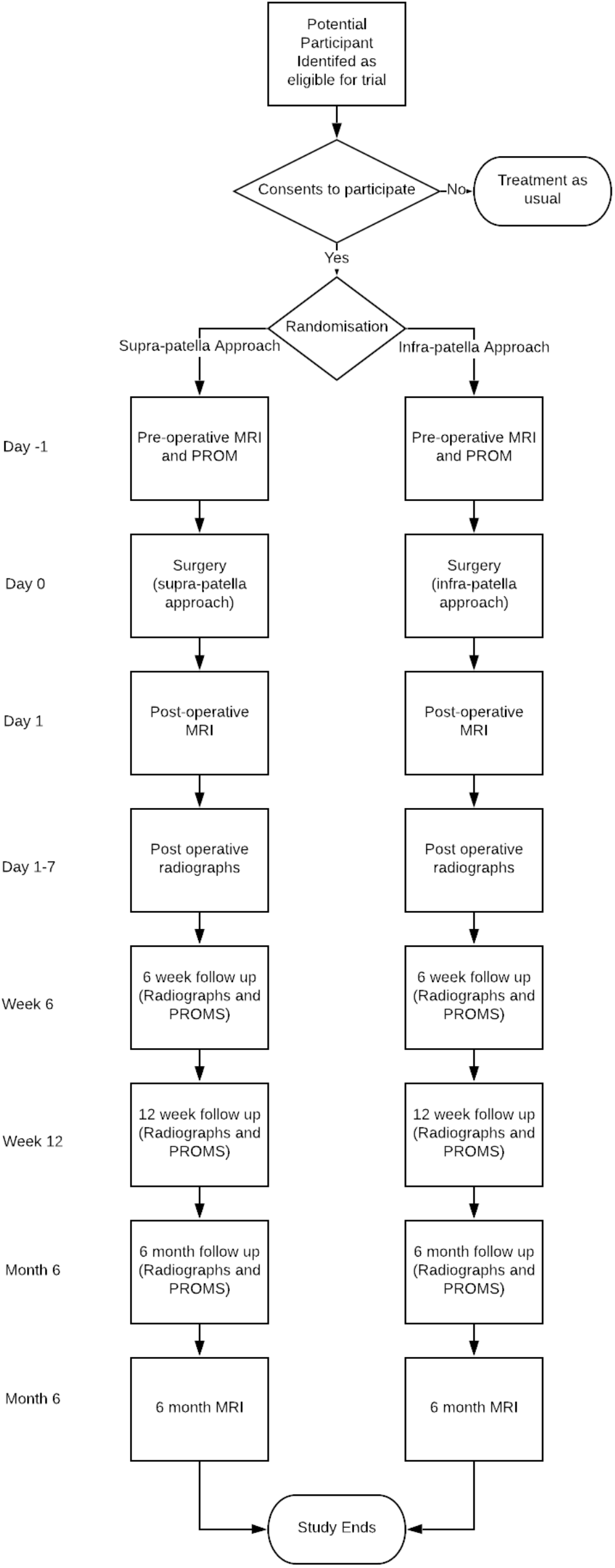
Study Participant Flow Diagram.

### 4.1 Study Assessments/Interventions

See Appendix 1 for an outline of when assessments and interventions occur.

#### 4.2.1 MRI Scans

Patients will undergo MRI scans of the knee of the affected leg at the following timepoints:

- Day -1 (the day before surgery)
- Day 1 (the day after surgery)
- Month 6

The MRI scan will include the following sequences:

- Axial, coronal, and sagittal PD FS
- Coronal PD
- Quantitative DESS (includes automatic in-line T2 map)

The estimate total scan duration is 30 minutes. Review of previous MRI scans has confirmed that the presence of the nail will not cause excessive metal artefact^9^.

Semiquantitative MRI analysis will be performed using the MRI osteoarthritis knee score (MOAKS) instrument ^10^. MOAKS cartilage grading is performed in a two-digit manner taking into account 1.) the percentage of area in each subregion affected by any cartilage damage and 2.) the percentage of area of each subregion affected by full thickness cartilage damage (both scored from 0 to 3; 0 = none, 1 = <10%, 2 = 10-75%, 3 = >75%). All knee joint regions are scored including the patellofemoral joint.

Quantitative MRI analysis will be performed using 3D-Cartilage Surface Mapping which enables integrated assessment of both cartilage thickness and T2 relaxometry^11^. T2 relaxation time is affected by cartilage water content and organisation of the collagen lattice and is an accepted biomarker of cartilage quality ^12^. All joint regions are analysed including the patellofemoral joint. While the original validation paper did not include retropatellar cartilage due to the characteristics of the included clinical cohort, the technique has been successfully applied to the retropatellar cartilage ^13^. Moreover, validation data for cartilage surface extraction are comparable to the femoral and tibial cartilage with mean distance error [95% limits of agreement] of 0.13 [-0.24, 0.50] mm (patella), 0.05 [-0.40, 0.50] mm (femur), 0.15 [-0.21, 0.52] mm (medial tibia) and 0.11 [-0.64, 0.85] mm (lateral tibia).

#### 4.1.2 PROMS

Patients will be asked to complete the following questionnaires pre-operatively and at each outpatient appointment:

- Lysholm Knee Score
- Tegner Activity Questionnaire
- EQ-5D-5L

#### 4.1.3 Clinical Outcome Data

We will also collect the following information for each case:

- Basic demographic information
- Operative information (time, blood loss, total radiation exposure)
- Post-operative complications (including infection, venous thromboembolism, compartment syndrome).
- Time to union
- Return to work/employment status

### 4.2 Definition of End of Study

The end of study is the date of the 6 month follow up appointment and MRI of the last participant.

## 5 STUDY SETTING

The study will be conducted at CUHNHSFT, Cambridge. This is a Major Trauma Centre serving the East of England geographical region.

## 6 SAMPLE AND RECRUITMENT

### 6.1 Eligibility Criteria

Participants will be drawn from patients admitted to CUHNHSFT under the Trauma and Orthopaedic Service.

#### 6.1.1 Inclusion criteria

- Age 18 – 50
- Male or female
- Closed tibial shaft fracture suitable for intramedullary nailing
- Able to consent
- Able to undergo MRI scanning
- Able to attend follow up for 6 months post operatively
- Suitable for surgery

#### 6.1.2 Exclusion criteria

- Polytrauma
- Ipsilateral acute ligamentous knee injury
- A contra-indication to MRI scanning (for example certain internal cardiac defibrillators)

### 6.2 Sampling

All individuals who meet the inclusion criteria will be approached to take part in the study from the time of study commencement until recruitment has completed (20 individuals). If they decide to take part in the study they will be randomised to one form of treatment. Participants will be randomized using the sealed envelope online randomization service (www.sealedenvelope.com). “A” will indicate supra-patella nailing. “B” will indicate infra-patella nailing.

#### 6.2.1 Size of sample

The novel nature of this study means that no previous data is available to enable us to perform a precise power calculation. However, we have used the data in a previous study^*13*^ to provide a reasonable estimation of the sample size required. This study looked at the effect of exercise on knee cartilage, including the patella. Taking the T1rho change from this paper a power calculation was performed using G*Power 3.1 with the following criteria:

- ANOVA, repeated measures (3), between factors (2)
- 0.8 power
- Type 1 error rate 5%

This produces a total sample size of 14, with an actual power of 0.86. Using a total sample size of 20 will allow us to accommodate for potential loss to follow up and any potential underestimation of the standard deviation between groups taken from the study mentioned.

As this is a pilot study the information obtained will allow us to perform precise power calculations for a potential future multi-centre study involving Major Trauma Centres nationwide.

#### 6.2.2 Sampling technique

All eligible individuals will be approached.

### 6.3 Recruitment

Potential participants will be initially identified and approached by members of the direct care team.

#### 6.3.1 Sample identification

Individuals admitted to CUHNHSFT with a tibial fracture will form the population from which potential participants are approached.

#### 6.3.2 Consent

Potential participants will be provided with an information sheet and will have an opportunity to discuss the study with a member of the study team and ask any questions. If they elect to take part, they will be asked to sign a consent form which will be kept on record in the trial documentation.

## 7 ETHICAL AND REGULATORY CONSIDERATIONS

The only study interventions that do not form part of standard of care are the completion of patient reported outcome measures (PROMS) and MRI scanning. MRI scanning is widely regarded as a safe procedure and does not involve any exposure to ionising radiation. Individual’s care pathway will not be disrupted by their decision to take part and as such they will not be disadvantaged by taking part.

### 7.1 Assessment and management of risk

There are no significant risks to taking part over and above those experienced as part of the normal standard of care (undergoing surgery and standard of care radiographs). As such, there is not felt to be any significant additional risk from taking part in the study. The implants used in the study are safe to be exposed to an MRI magnetic field in the immediate post-operative period.

### 7.2 Research Ethics Committee (REC) and other Regulatory review & reports

A favourable ethical opinion has been obtained from the East of England – Cambridge South Research Ethics Committee (reference 21/EE/0178). Approval has also been received from the NHS Health Research Authority.

Substantial amendments that require review by NHS REC will not be implemented until that review is in place and other mechanisms are in place to implement at site.

All correspondence with the REC will be retained.

It is the Chief Investigator’s responsibility to produce the annual reports as required. The Chief Investigator will notify the REC of the end of the study.

An annual progress report (APR) will be submitted to the REC within 30 days of the anniversary date on which the favourable opinion was given, and annually until the study is declared ended.

If the study is ended prematurely, the Chief Investigator will notify the REC, including the reasons for the premature termination.

Within one year after the end of the study, the Chief Investigator will submit a final report with the results, including any publications/abstracts, to the REC.

### Regulatory Review & Compliance

Before the study enrols patients into the study, the Chief Investigator or designee will ensure that appropriate approvals from Cambridge University Hospitals NHS Foundation Trust are in place. Specific arrangements on how to gain approval from participating organisations are in place and comply with the relevant guidance.

For any amendment to the study, the Chief Investigator or designee, in agreement with the sponsor will submit information to the appropriate body in order for them to issue approval for the amendment. The Chief Investigator or designee will work with sites (R&D departments at NHS sites as well as the study delivery team) so they can put the necessary arrangements in place to implement the amendment to confirm their support for the study as <u>amended</u>.

### Amendments

Amendments will be initially passed to the Research & Development Department at Cambridge University Hospitals NHS Foundation Trust. Following their approval they will be sent to the appropriate NHS REC for formal review.

No amendment will be put into place until approved by the NHS REC (or confirmation of receipt received in the case of minor amendments).

Amendment history will be recorded in the protocol. Old versions (including of associated paperwork) will be archived to prevent any ongoing use of superseded paperwork.

### 7.3 Peer review

Peer review has been undertaken by the Research Advisory Committee of Addenbrooke’s Charitable Trust.

### 7.4 Patient & Public Involvement

Patient and Public involvement in study design and design of associated paperwork has been undertaken with the Cambridge University Hospitals Patient and Public Involvement Panel.

### 7.5 Protocol compliance

Protocol compliance will be monitored by the data monitoring committee. All deviations will be reported to this committee. Significant deviations or breaches will be reported to the Sponsor and REC as necessary.

#### 7.5.1 Definition of Serious Adverse Events

The additional study procedures (MRI scanning and completion of outcome measures) are not believed to pose a risk of causing a serious adverse event.

However, the normal treatment that participants will be receiving (surgical procedures involving general anaesthetic and/or spinal anaesthesia) does carry a risk of harm. As such, any event which:

- Results in death
- Is life threatening
- Significantly extends the inpatient stay
- Or causes significant excess morbidity

will be treated as an SAE and managed accordingly.

#### 7.5.2 Reporting Procedures for Serious Adverse Events

All SAEs will be reported to the REC, as well as the study sponsor, as required.

### 7.6 Data protection and patient confidentiality

Identifiable patient information will be held on NHS systems (PACS and EPIC computerised records system).

- Study data will be anonymised at the earliest opportunity for the purposes of ongoing analysis. It will be stored in a linked-anonymous format (i.e. a participant code will be allocated to each individual and a separate cross-reference table will be held securely to link this code to the individual’s identity. Only members of the direct care team will have access to this link table.
- Clinical data held on NHS systems will not be deleted as it will form part of the patient record.
- Linked-anonymous data will be deleted after 15 years.
- The data custodian will be the chief investigator.
- All data handling, processing, and storage will be in line with GDPR and the Data Protection Act 2018.

### 7.7 Indemnity

Indemnity will be provided by the NHS, as the primary sponsor of the study. This indemnity will cover all aspects of study conduct, design, and management.

### 7.8 Access to the final study dataset

All study group members will have access to the full dataset. Only direct care team members will have access to patient identifiable information.

## 8 DISSEMINIATION POLICY

### 8.1 Dissemination policy

The data produced will be owned by Mr Andrew Carrothers as the data controller.

A final report will be produced and held by the R&D department of Cambridge Hospitals NHS Foundation Trust.

The final data will be used to produce a manuscript for publication and dissemination at professional conferences. All investigators will be acknowledged in these publications.

Funders will be acknowledged in all publications but have no control over the timing or location of publications.

On enrolment, participants will be advised of the location of the publication of results (funder website).

Participant level data will not be published. The protocol will be uploaded to ClinicalTrials.gov at the completion of the study in line with their guidelines.

## Data Availability

No data is available as this is only the study protocol.

## Disclaimer

This publication presents independent research partially funded by the National Institute for Health Research (NIHR). The views expressed are those of the author(s) and not necessarily those of the NHS, the NIHR or the Department of Health and Social Care.

## 10. APPENDICIES

### 10.1 Appendix 1 – Schedule of Procedures

| Time | Procedure |  |  |  |  |  |
| --- | --- | --- | --- | --- | --- | --- |
|  | Informed Consent | MRI | Surgery | PROM | Radiographs | Outpatient Appointment |
| On Admission | X |  |  |  | X* |  |
| Day Admission to -1 |  | X |  | X |  |  |
| Day 0 |  |  | X |  | X |  |
| Day 1 |  | X |  |  |  |  |
| Day 1-7 |  |  |  |  | X |  |
| Week 6 |  |  |  | X | X | X |
| Week 12 |  |  |  | X | X | X |
| Month 6 |  | X |  | X | X | X |

Events in green are normal standard of care events that would take place whether or not the patient was taking part in the trial.
\* This exposure would be on attendance to the Emergency Department to diagnose the injury and so will take place prior to consideration for enrolment.

## FUNDER

Addenbrooke’s Charitable Trust Addenbrooke’s Hospital Hills Road, Cambridge CB2 0QQ

## FINANCIAL AND NON FINANCIALSUPPORT GIVEN

£24,000 to support cost of MRI scanning

## ROLE OF STUDY SPONSOR AND FUNDER

The review committee of the funder provided an internal peer review of the study protocol. Apart from this the funder has no role in study design, conduct, data analysis and interpretation, manuscript writing, and dissemination of results.

The sponsor has no role in study design, conduct, data analysis and interpretation, manuscript writing, and dissemination of results.

## STUDY SUMMARY

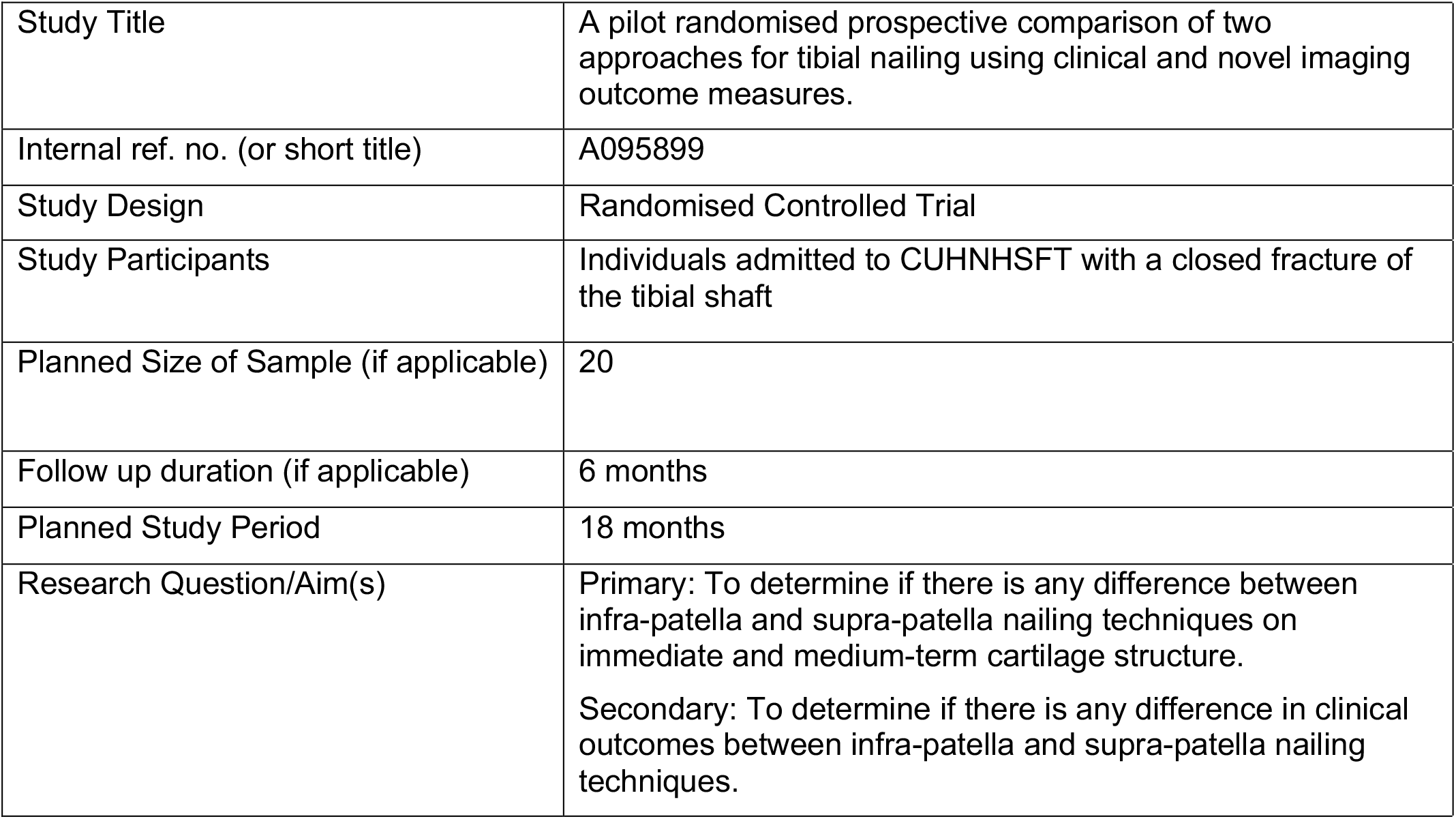

|  |  |
| --- | --- |
| Study Title | A pilot randomised prospective comparison of two approaches for tibial nailing using clinical and novel imaging outcome measures. |
| Internal ref. no. (or short title) | A095899 |
| Study Design | Randomised Controlled Trial |
| Study Participants | Individuals admitted to CUHNHSFT with a closed fracture of the tibial shaft |
| Planned Size of Sample (if applicable) | 20 |
| Follow up duration (if applicable) | 6 months |
| Planned Study Period | 18 months |
| Research Question/Aim(s) | Primary: To determine if there is any difference between infra-patella and supra-patella nailing techniques on immediate and medium-term cartilage structure.<br>Secondary: To determine if there is any difference in clinical outcomes between infra-patella and supra-patella nailing techniques. |

## ROLES AND RESPONSIBILITIES OF STUDY MANAGEMENT COMMITEES/GROUPS & INDIVIDUALS

### Internal Data and Safety Monitoring Committee

This committee is composed of Mr Andrew Carrothers (as PI), Prof Andrew McCaskie (as an orthopaedic surgeon who will not be conducting any of the surgery), and Dr James MacKay (Radiologist) and will oversee data and safety issues during the conduct of the trial.

### Patient and Public Involvement

Patient and Public Involvement was sought at the study design and study paperwork development stage. This was provided by Cambridge University Hospitals NHS Foundation Trust Patient and Public Involvement Panel.

## Notes

All authors declare funding from Addenbrooke’s Charitable Trust to support this project.

The Department of Trauma and Orthopaedics at Addenbrooke’s Hospital receives funding from Depuy Synthes to support a Trauma Fellow post.

### Competing Interest Statement

All authors declare funding from Addenbrooke's Charitable Trust to support this project.
The Department of Trauma and Orthopaedics at Addenbrooke's Hospital receives funding from Depuy Synthes to support a Trauma Fellow post.
BMD reports funding from an NIHR Clinical Lectureship, not directly related to this study.
EA has nothing else to declare.
DC has nothing else to declare.
PH has nothing else to declare.
JW has nothing else to declare.
JMcK reports grants and/or consultancy payments from GlaxoSmithKline, Moximed, GE Healthcare, and employment by AstraZeneca, none directly related to this study.
AMcC reports work for Taylor and Francis (publishers), membership of an advisory board for Arthritis Research UK, and a patent for cartilage repair unrelated to this study.
AC reports grants and/or consultancy payments from NIHR, Stryker, Matortho, Zimmer Biomet, as well as income from expert testimony, none directly related to this study.

### Clinical Trial

NCT04831671

### Funding Statement

This study was funded by Addenbrooke's Charitable Trust.

### Author Declarations

East of England Cambridge South Research Ethics Committee (reference 21/EE/0178) of the NHS gave ethical approval for this work.

